# A longitudinal feasibility study on fostering daily conversation habits centered on perspective taking and time orientations for cognitive health: Protocol for a single-arm intervention in Japanese local communities

**DOI:** 10.1101/2025.08.22.25334249

**Authors:** Takuya Sekiguchi, Kazumi Kumagai, Hikaru Sugimoto, Mihoko Otake-Matsuura

## Abstract

**Background:** Dementia prevention has the potential to bring considerable benefits in diverse contexts. Conversation requires various cognitive functions and is therefore considered a natural multi-component preventive measure that can be practiced in daily life. Although an emerging line of research supports the potential of conversation-based cognitive interventions, further research is needed to elucidate which elements are effective and through what mechanisms. In addition, given that previous studies have shown that older adults embedded in social networks with a high proportion of non-family members have higher cognitive function, fostering social interactions between older adults with those beyond their family would be beneficial, while assessing the ecological validity of delivering conversation-based program in naturalistic community settings is essential. Furthermore, it remains to be clarified whether participants can integrate lessons from conversation-based programs into their daily routines in a voluntary and sustained manner. This study aims to examine the feasibility of the long-term conversation-based program, with preliminary analyses of its key components for future rigorous effectiveness tests.

**Methods:** The study is a single-arm longitudinal intervention in a community setting in Japan. Participants will attend monthly sessions over five years, engaging in conversations based on the Coimagination method. The feasibility and adherence are assessed mainly by dropout rates and speech frequencies in sessions, respectively. The primary outcome is verbal fluency, repeatedly improved by previous conversational interventions. The key components of the intervention such as perspective taking and time orientation will also be measured. Preliminary intervention effects will be assessed by the interaction between adherence and time in a mixed-effects model.

**Discussion:** The study following this protocol is expected to provide insights into the feasibility and ecological validity of our conversation-based program and its barriers, as well as preliminary findings for conducting a future long-term randomized controlled trial to rigorously verify the intervention effects. Because the intervention will be conducted in parallel in two regions, suggestions for external validity are also expected.

**Trial registration:** Prospectively registered in the UMIN Clinical Trials Registry (UMIN000057738).

## 1 Introduction

### 1.1 Background

In societies with a growing proportion of older adults—Japan being a typical example [1] and the setting of this study—preventing dementia is of great significance to maintain the quality of life of older adults themselves and their caregivers [2], as well as reducing the economic burden at the national level [3], [4].

Dementia can cause a wide variety of symptoms, including personality changes and cognitive decline. Alzheimer’s disease is the most prevalent subtype and its hallmark early symptom is progressive memory impairment, e.g., difficulty remembering recent events [5]. Developing measures aimed at maintaining broader cognitive functions as well as memory, and preventing or delaying the onset of dementia, is an urgent issue.

Findings on strategies for improving the performance of cognitive tasks would seem to offer an effective solution to this issue. However, even if one performs well on a specific task, it is unclear whether this improves his/her general cognitive function necessary for daily life [6], [7]. Therefore, a multi-component approach, which involves applying multiple activities that may be beneficial for maintaining cognitive function in a complex manner to daily life, is considered promising [6], [8], [9].

Social interaction requires complex cognitive processes and is therefore compatible with the multi-component intervention approach. In fact, it has been consistently reported to be associated with improved cognitive function [10]. However, should social interactions be considered the groundwork of multi-component interventions, a crucial point to address is understanding which aspects of social interaction matter most, with whom these interactions occur, and the mechanism behind the association. References [11], [12] are suggestive observational studies in this sense, reporting better episodic memory in people embedded in social networks with a high proportion of non-family members. While Ref. [11] offers one possible interpretation of this result that more active communication is necessary to maintain emotional closeness in friendships than in family, scrutinizing the contents of conversations with non-family persons may provide insight into which types of contents are cognitively stimulating.

In this context, conversation, one of the fundamental elements of daily social interactions, has attracted attention as a promising intervention component. A randomized controlled trial (RCT) [13] conducted a web-based intervention, in which socially isolated older adults experienced a semi-structured conversation with interviewers, and observed the positive intervention effect on global cognition. This line of research suggests that conversation can take advantage of its everyday nature to transform daily life into a natural and sustainable effective intervention program. To achieve this, refining how people engage in conversation is a key challenge because not all types of conversation could be equally effective in maintaining cognitive function.

The Coimagination method is a conversation-based multi-component cognitive intervention program [14] and has been the subject of a series of evaluation studies conducted with a focus on these considerations. This program involves structured group conversations. Participants take photos related to a predetermined conversation topic in advance, and on the day of the session, they explain their photos and ask questions about others’ photos. The presentation and Q&A times are strictly monitored by the system, requiring participants to communicate information efficiently within tight time constraints. To perform this, multiple cognitive functions are required, including planning, attention, and working memory, as well as word retrieval ability.

In an RCT conducted to verify the efficacy of the Coimagination session [15], a positive intervention effect was observed on letter fluency. Given the letter fluency test requires generating words under time constraints while avoiding repetition, which overlaps with the cognitive demands of the Comagination sessions, this result is theoretically plausible.

Specific findings have also been obtained, indicating the promise of components that constitute the Coimagination method and their potential to enhance its intervention effects. The need to attend to others’ view is further intensified by the requirement to ask questions about other members’ presentations. The result of [16] is suggestive in this regard: namely, those who tend to compare themselves to others from the perspective of other persons’ opinions rather than from the perspective of other persons’ abilities had a higher global cognitive function. As this study is cross-sectional, no definitive conclusions can be drawn about causality. However, it is worth conducting an intervention study to examine whether cognitive function improves through conversations with various types of people and the experience of taking the perspective of others.

Another feature of the Coimagination method is that photographs used in the conversation are required to be taken in the near past from the session. This was designed with the aim of avoiding the loss of episodic memory before recent information is integrated through conversation. In a sense, this method can be seen as one designed to guide participants’ time orientation, i.e., cognitive emphasis on specific time, toward the recent past. Related to this, Ref. [17] analyzed transcription data from unstructured group conversations of older adults and found that those who scored higher on a pre-measured logical memory task had more utterances whose content was characterized by recent knowledge. This study also requires caution about causality; that is, this result alone does not necessarily indicate that talking about recent events leads to the maintenance of cognitive function. Nevertheless, it is reasonable to use this finding as motivation for intervention studies that focus on talking about recent events.

The RCTs on the Coimagination method left several unanswered questions due to their design. One key issue is the long-term effects of the intervention. The RCTs [15] and [18] are designed to evaluate the intervention effect of face-to-face Coimagination-based conversation sessions. For these studies, a three-year follow-up was preregistered to investigate the impact of the intervention on participants’ daily lives afterward, and data cleaning and analysis are currently underway. However, participants in the intervention group of these studies only engaged in the program for three months, and subsequently only underwent assessments during follow-up. The effects of continued intervention over several years on cognitive function have not been verified.

Closely related is the question of how key elements of the Coimagination-based intervention, such as perspective taking and time orientation, evolve over time and how these changes influence the magnitude of intervention effects. An RCT prospectively registered in [19], recently completed but pending analysis, was designed to explore the relationship between these elements and the intervention effect using psychological scales. However, that intervention study was conducted via a remote application and has not verified the effects in face-to-face settings. Furthermore, that study involves an intervention lasting approximately six months. Therefore, the long-term transformation process of these elements remains unclear.

An overarching unresolved issue is the feasibility and ecological validity of the long-term community-based conversation intervention program. The aforementioned RCTs were of short duration and included participant incentives. Possibly due to these factors, there were no dropouts in [15], [18]. However, to assess whether the lessons learned from the multi-component cognitive intervention program can be applied in everyday community life, an exploratory feasibility and ecological validity study is needed that examines whether community members can voluntarily build the necessary environment and operate equipment for participation without financial incentives. Furthermore, because previous RCTs have not implemented the same program in different regions in parallel, the external validity in terms of regional differences has not been verified.

### 1.2 Objectives

In light of this background, this paper provides a protocol for conducting preliminary research for future rigorous studies to clarify the following issues. The first issue is the long-term effects of an intervention program primarily consisting of the Coimagination method on cognitive function. We have set verbal fluency as the primary outcome. The reasons are, first, that the Coimagination session and verbal fluency tests are similar in requiring participants to produce language in a limited time, and they are considered to involve common abilities such as word retrieval and executive function [20], [21]; and second, that verbal fluency has been repeatedly shown to improve in conversation-based intervention studies [15], [22], and this study allows us to examine the extent of improvement achieved through a longer-term intervention compared to those studies.

The second issue is how specific components of the program impact the outcomes. Participants are required to acquire other participants’ perspectives and retrieve recent episodes in order to properly perform the Coimagination session, which can be considered an activity related to logical memory. To examine whether these factors contribute to the maintenance and improvement of verbal fluency and logical memory, we will also focus on social comparison orientation and temporal orientation as secondary outcomes.

The third issue is the prospects for sustained implementation. Our future goal is to develop multi-domain intervention programs that incorporate the Coimagination method as a key element in various communities. Thus, this study also aims to assess the long-term feasibility and ecological validity of the program.

In summary, the purpose of this study is to explore the feasibility of fostering daily conversation habits centered on perspective taking and time orientations via long-term Coimagination-based intervention in multiple regions, and preliminary evaluation of its effect on multiple cognitive domains.

## 2 Methods and design

### 2.1 Study design

This protocol describes a single-arm (i.e., without a comparator group) longitudinal feasibility study with exploratory evaluation of cognitive outcomes in the group conversation-based intervention. Participants will have face-to-face group conversations once a month. Primary and secondary outcomes will be basically evaluated at baseline, annually during the intervention, and at endpoint. The intervention period is planned to be five years, but is subject to change, depending on human and financial resources. The SPIRIT flow diagram of this study is shown in Table 1.

**Table 1:** The SPIRIT flow table for the single-arm feasibility study.

| **Trial Phase** | **Enrollment** | **Baseline** | **Intervention** | **Endpoint** |
| --- | --- | --- | --- | --- |
| **Enrollment** |  |  |  |  |
| Session demonstration | ✓ |  |  |  |
| Informed consent | ✓ |  |  |  |
| **Intervention** |  |  |  |  |
| Practice session |  |  | ✓ |  |
| Intervention |  |  | ✓ |  |
| **Assessments** |  |  |  |  |
| **Background** |  |  |  |  |
| Basic characteristics |  | ✓ |  |  |
| Kihon checklist |  | ✓ | ✓ | ✓ |
| **Implementation** |  |  |  |  |
| Number of questions |  |  | ✓ |  |
| Dropout rate |  |  | ✓ |  |
| **Cognition** |  |  |  |  |
| Verbal fluency |  | ✓ | ✓ | ✓ |
| LMs I & II |  | ✓ | ✓ | ✓ |
| MMSE-J |  | ✓ |  | ✓ |
| MoCA-J |  | ✓ | ✓ | ✓ |
| NCGG-FAT |  | ✓ | ✓ | ✓ |
| **Psychometrics** |  |  |  |  |
| SCO |  | ✓ | ✓ | ✓ |
| IRI |  | ✓ | ✓ | ✓ |
| ZTPI |  | ✓ | ✓ | ✓ |
| PVQ-II |  | ✓ |  | ✓ |
| NEO-FFI |  |  | ✓ | ✓ |
| LSNS-6 |  | ✓ | ✓ | ✓ |
| UCLA-LS3 |  | ✓ | ✓ | ✓ |
| WHO-5 |  | ✓ | ✓ | ✓ |
| **Others** |  |  |  |  |
| Blood biomarkers |  |  | ✓ | ✓ |
| Conversation data |  |  | ✓ |  |

### 2.2 Study areas

The intervention will be conducted in two Japanese cities. Wako, located in Saitama Prefecture (which borders Tokyo in eastern Japan), has a population of 84,890, with approximately 18.1% aged 65 or older as of January 2025 [23]. Kishiwada, located in Osaka Prefecture (which is the third most populous prefecture in Japan and is located in the western part of the country), has a population of around 185,870, with approximately 28.5% aged 65 or older as of July 2025 [24]. Given that the percentage of people aged 65 and over in Japan’s total population was 29.3% as of February 2025 [25], Wako has the lower ratio, while Kishiwada is on par with the national average.

To conduct this study, we have already entered into joint research agreements with the municipal governments of both cities. For Kishiwada, the agreement included Nobana Health Promote Co., Ltd., forming a three-party collaboration. These partnerships currently support participant recruitment and are expected to support logistical aspects of the study, such as providing venues for intervention sessions.

### 2.3 Eligibility criteria

Our goal, looking beyond this study, is for social interactions based on group conversations tuned toward maintaining cognitive function to take root in the community. Overly strict eligibility criteria can lead to underestimating operational barriers and are inadequate for verifying the feasibility of our goal.

Therefore, this study sets the following moderate inclusion criteria: (1) aged 18 years of age or older, which is the legal definition of adulthood in Japan, (2) able to provide written informed consent, (3) available to participate in face-to-face group conversations once a month on a designated date and time, and (4) able and willing to undergo study outcome assessments on a designated date and time. The reason for setting (1) was that, with community implementation in mind, we judged that younger individuals should not be strictly excluded from participation.

The exclusion criteria are as follows: (1) having difficulty participating due to serious illness, (2) being unwilling to speak and only listen to others, and (3) unable to prepare the equipment and environment necessary to participate in the group conversation.

### 2.4 Recruitment strategy

In Kishiwada, a public announcement was made on May 1, 2025, regarding the date and time of the briefing session for those considering participating in this study. Therefore, this date is defined as the date on which recruitment for this study began. Approximately 100,000 flyers were distributed and the related article was also published on the city’s official website on the same day. The announcement stated that this study aimed to prevent dementia through conversation and that it was a joint research project between the city and RIKEN. A public announcement of the same information was also released on June 1, 2025. The briefing sessions have already been held in a civic center in Kishiwada. Originally, we announced that the briefing session would be held on June 3, 2025. However, in response to requests from potential participants, we arranged an additional briefing session on June 19, 2025. Applications were accepted from June 3 until June 25, 2025. Originally, if there had been a high number of applicants, a lottery among the participants was planned to be held. However, because we had not yet reached the target sample size, after accepting all the applications, we also accepted additional individuals who expressed interest in the present study. On July 8 and 9, 2025, trial sessions were held for those interested in participating in conversation sessions. At the time of writing this manuscript, approximately forty participants are expected to attend the intervention. Consent acquisition and baseline assessments began on August 6, 2025, and were completed by the 21st. The first intervention session is scheduled for early September and will primarily serve as a practice run.

In Wako, as in Kishiwada, the a public announcement with the same information was released on May 1, 2025. The same content has been published in the public announcement issued on June 1, July 1, and August 1, 2025. The briefing session was held on July 2, 2025, in a civic center in Wako. Different from Kishiwada, in Wako, the briefing session was preceded by a debriefing meeting on the result of the research [26], [27] previously conducted in collaboration with the city. Information about the briefing session and the intervention program was also posted on the web application that features community events, GBER [28]. On July 22, 2025, the trial session was held for those interested in participating in conversation sessions. For the same reason as Kishiwada, we arranged an additional briefing session on August 25, 2025. Applications will be accepted from that day until August 27, 2025. If there are many applicants, a lottery will be held to select participants on a day between late August and early September. The first intervention (rehearsal session) is scheduled for mid-October.

### 2.5 Interventions

We use the same system as was used in the RCTs of the Coimagination based-intervention program [15], [18]. Schematic of the program and its system are shown in Figure 2 in [15]. Participants will download an application developed by our team specifically for the conversation sessions based on the Coimagination method onto their smartphones in advance. Although participants are expected to use their own smartphones, we have a few available for loan. Currently, our app is only compatible with Android devices. Therefore, if participants are using other types of devices or if their devices are outdated and cannot run the app properly, we will lend them our smartphones.

The conversation topic will be announced to participants about a week before each session. Our app has a camera function, and participants are instructed to take as many photos as possible using the app that are related to the topic. Before the conversation session begins, participants will select two of the photos they have taken and upload them to the web system. This is the preparation phase for each session. (The schematic of the app in the whole system is shown in Figure 2A in [15].)

The face-to-face session day consists mainly of the moderated phase and the recall phase.

The flow for the day is as follows:

(1) Icebreakers such as asking participants about their experiences over the past month
(2) Study sessions on topics such as the effects of social interaction, including conversation, and lifestyle on health
(3) Review points to keep in mind when engaging in the Coimagination method, such as talking about recent events and asking questions
(4) Group conversation based on the Coimagination method (Moderated conversation phase)
(5) Participants write down their recollections of the session, such as who said what (Recall phase)
(6) Participants hold a simple tea time for chatting
(7) Participants are asked to write a brief description of about 200 characters on our app for each of their photos used in the session, summarizing what they talked about

Sessions will be held once a month for approximately two hours.

Here, the main component of this study, the moderated conversation phase (4), should be supplemented. (A block diagram of the moderated conversation phase in the program is shown in Figure 4 in [15].) The first participant presents two photos, spending one minute per photo (a total of two minutes). For each photo, that participant first describes when and where it was taken. All participants do this sequentially. After all presentations are finished, a two-minute Q&A session per photo (total of four minutes per participant) is conducted, also sequentially for all participants. In the Q&A session, participants are instructed to focus specifically on generating questions rather than simply sharing their impressions of the photos. This is the modified point from previous RCTs to promote perspective taking via conversation. All timing is controlled by a conversational robot developed by our team. The robot cues the start and end of conversations and designates speakers. Instructions given by the robot are based on our previous RCT (see Table 2 in [15]).

Conversation groups are formed by prioritizing an approximately gender balance within group, accommodating participants’ preferred schedules, and making efforts to have friends or family members participate on the same day. Other assignments are made based on the order of the name list. Each group consists of about ten participants, which is then divided into two subgroups of five members each. This partition is not fixed and may change during the intervention period.

This study aims to verify whether the conversational style pursued by the Coimagination method is feasible in everyday life, particularly in terms of its compatibility with fostering social interaction. Therefore, rather than simply conducting structured conversation sessions (4), we also conduct conversations that are closer to natural conversations, such as (1) and (6), to promote interaction among participants. In addition, through (2) and (3), we will raise participants’ motivation and ensure that they do not overlook the aims of our intervention. Participants may learn the importance of perspective taking and time orientation. Instructors may explicitly recommend participants to ask questions by imagining the perspective of the speaker in order to enhance perspective taking. Instructors may also explicitly recommend participants to talk about recent topics in order to orient time towards recent and present. These instructions are done for the group conversation based on the Coimagination method (4), as well as ice breaking (1) and tea time (6). Participants are asked to exercise asking questions and taking about recent topics during daily conversations even outside of the sessions. The purpose of these instructions is to promote cognitively engaging conversation habit. This is intended to enhance the effectiveness of the intervention despite its low frequency of once a month.

Furthermore, by conducting (5) and (7), we aim to assist participants in retaining the most recent episodes obtained in the conversation session and to foster their perspective taking. Participants will not only listen to the conversation sessions of the five-member subgroup to which they belong but also observe all members of the other subgroup. They will then recall and describe the speech contents of those participants as well during (5).

This program is still in the process of refinement to improve its effectiveness. We consider this study exploratory, also serving as a platform for refining the intervention methods through trial and error. In particular, how the robot engages in the conversations is one of our major concerns.

This study is interested in whether such activities can take root in the community. To eliminate the effect of monetary incentives, no remuneration will be provided to participants.

### 2.6 Measures

#### 2.6.1 Background information

As basic characteristics, our questionnaire will ask participants about age, sex, usual health status, cohabitants, educational background, frequency of daily device usage, subjective assessment of household financial situation, and subjective well-being. Additionally, functional status will be assessed by using the Kihon checklist [29], [30].

#### 2.6.2 Implementation outcomes

Although this study adopts a single-arm design, we should obtain data illuminating the mechanism of intervention effects for future full-scale research. For that purpose, in this study, participants are allowed flexibility in engagement in the intervention program, and the degree of involvement is quantified to investigate the effects of the intervention program on cognitive function. Specifically, we will use the number of questions during the session as a proxy for adherence. This will be counted using the conversation data described in a later section. The dropout rate is also recorded as an indicator of acceptability.

#### 2.6.3 Primary intervention outcome

In multi-component intervention studies, which elements of the intervention affect the outcome is often unclear. If we set the global cognition score as the outcome, the driving factor for the intervention effect is even more elusive. As mentioned in Section 1.2, considering the overlap with the abilities required by the Coimagination session, we have selected verbal fluency as the primary outcome.

The verbal fluency test has two types: letter fluency and category fluency tests. The former (the latter) is a task that requires participants to generate as many words as possible within one minute that begin with a specific letter (that belong to a specific category). The number of words generated will be directly considered as the score for this task.

#### 2.6.4 Secondary intervention outcomes

This study includes the following variables as secondary outcomes to deepen our understanding of the intervention’s nature:

- Social Comparison Orientation Scale (SCO) [31], [32]
- Interpersonal Reactivity Index (IRI) [33], [34]
- Zimbardo Time Perspective Inventory (ZTPI) [35], [36]
- Lubben Social Network Scale (LSNS-6) [37], [38]
- University of California Los Angeles Loneliness Scale (UCLA-LS3 (SF-10)) [39], [40]
- World Health Organization-Five Well-Being Index (WHO-5) [41], [42]
- Personal Values Questionnaire II (PVQ-II) [43]
- NEO Five Factor Inventory (NEO-FFI) [44]
- Logical Memory I and II (LMs I & II) in the Japanese version of the Wechsler Memory Scale-Revised [45]
- Japanese version of the Mini-Mental State Examination (MMSE-J) [46], [47]
- Japanese version of the Montreal Cognitive Assessment (MoCA-J) [48], [49]
- National Center for Geriatric and Gerontology-Functional Assessment Tool (NCGG-FAT) [50]

SCO has two sub-scales that measure the tendency to compare oneself with others in terms of ability and in terms of opinion. As mentioned in the Introduction section, Ref.[16] showed a positive cross-sectional correlation between the opinion subscale score and the MoCA score. This longitudinal study will examine whether participants’ opinion scale scores increase over time and whether this trend is greater among those with a higher level of commitment to the program. Whether the increase is in tandem with the increase in cognitive function will also be examined. IRI is a multi-dimensional measure of empathy and was included in the outcome with the same motivation as SCO, especially since it includes perspective taking as a subscale.

ZTPI is a measure of attitude toward time (past, present, and future). The Coimagination method aims to encourage older adults to retain recent episodes. We examine how the measured time orientation changes in response to the Coimagination-based intervention, which directs attention to a specific time (i.e., the recent past) and whether it influences the magnitude of intervention effects.

Given that this intervention involves social interaction, it is expected to alleviate loneliness, maintain or improve mental health, and sustain or expand social networks. These outcomes will be assessed longitudinally using UCLA-LS3, WHO-5, and LSNS-6, respectively.

PVQ-II is a questionnaire that asks which life domains participants prioritize, the reasons why they value them, and the extent to which they live in accordance with their values. By combining this with the conversation data described in the next subsection, we could investigate the relationship between values and conversation content, as well as obtain basic information for personalizing conversation topics. NEO-FFI will be used for the same purposes: namely, to investigate whether personality traits can change and how they relate to the content of conversations, with the goal of using these findings to refine our intervention program.

The Coimagination method aims to encourage older adults to retain recent episodes. We hypothesize that this will enhance their episodic memory. Thus, LMs I and II, which require participants to recall stories told by examiners, are included to measure this ability. Given that this study involves a multi-component intervention program, it is reasonable to expect improvements in global cognition. MMSE and MoCA-J are administered to verify this. NCGG-FAT, a tablet-based tool, is used to provide a more detailed assessment of multiple cognitive domains, including memory, attention, executive function, and processing speed.

Basically, the above outcomes are measured before and after the intervention, and annually during the intervention, as shown in Table 1. However, we judged that annual assessment by MMSE was unnecessary due to its limited sensitivity to detect cognitive changes compared to MoCA. Instead, MMSE will be administered only at pre– and post-intervention, to allow comparability with other longitudinal studies using MMSE. Moreover, considering that the PVQ-II is designed to assess core personal values that remain relatively stable over the lifespan and substantial outcome changes are unlikely, we plan to conduct it only pre– and post-intervention to minimize participant burden. Because NEO-FFI measures personality traits, which are generally considered to be stable, it will only be administered twice during the study period. Due to scheduling constraints, the assessments will not be strictly before and after the intervention, but will take place at two points during the intervention that are as close as possible to the baseline and the endpoint.

#### 2.6.5 Others

In addition to these established scales and assessment tools, participants’ conversations will be recorded and transcribed. In view of the preliminary nature of this study for future interventions, it is important to examine whether participants are engaged in our program as we intend and what conversation topics make them more likely to be so. Conversation data could be useful for this purpose.

Reference [17] is a typical example of such an approach. That study annotated the transcription data of conversations of older adults in the Coimagination session, which were collected from participants in the intervention group in [15], with labels indicating the time at which the event described in each utterance occurred. As a result, the percentage of sentences that refer to events in the past month varied depending on the session topic, despite instructing participants to share stories that are as recent as possible. Furthermore, using conversation data in the control groups (i.e., unstructured natural conversations) in [15], they found that those with better logical memory produced more sentences characterized by recent knowledge, and that such sentences were more often observed in the intervention conversation sessions with topics such as “Neighborhood landmarks” and “Found on a 10-minute walk”. Based on these results, and previous studies showing that walking is associated with a lower risk of dementia [51] and is a leisure activity that older adults easily initiate [52], they noted the prospect of an intervention in which the information gained while walking is output through conversation.

This data would also be used to investigate which conversational features are related to cognitive function. This study is a long-term intervention spanning five years, during which conversations on various topics will take place. Therefore, we may gain a deeper understanding of topics that are likely to elicit utterances with characteristics associated with high cognitive function. In addition, by integrating the conversation data with other data, we would build and refine the automated robot-moderation system to suit various conversation topics and diverse participant backgrounds.

We will also measure blood levels of amyloid beta and neurofilament light chain, which are indicators of neuronal health (higher levels suggest a worse condition). While we are interested in their longitudinal changes, we will also analyze the intervention effects conditioned on these biomarker levels. This is because, in Ref [18], our conversation-based program showed negative effects among individuals with high blood biomarker levels for some cognitive outcomes. Thus, this analysis will help us accurately capture both positive and negative aspects of our intervention. Blood biomarkers will be obtained once a year during the intervention and after its completion; however, the first draw will be scheduled, due to scheduling constraints, as close to the baseline as possible after the intervention begins.

#### 2.6.6 Timing of results dissemination

While the primary outcome will be analyzed and implementation outcomes will be reported after completion of the full intervention period, we plan to analyze selected secondary outcomes prior to the completion of the entire study and potentially published earlier. For example, due to the schedule constraints of the research grant (JSPS KAKENHI (Grant number 25K05587)) investigating longitudinal changes in time orientation among older adults using multi-modal data (such as questionnaires and conversations), which extends until March 2029, the analysis results of the ZTPI and conversation data would be published before the completion of this intervention. The hypothesis of that study is that this intervention, by encouraging conversations on recent events and participation in a long-term program, will foster a future-oriented attitude, leading to participants’ time orientation shifting from the past toward the present and future. However, reports related to the intervention effects will be published only after the intervention will have been completed.

### 2.7 Sample size and statistical analysis

We expect approximately 40 to 80 participants in Kishiwada and approximately 40 participants in Wako. These numbers were determined based on various practical constraints, such as budget, human resources, and availability of local facilities.

Due to the exploratory nature of this study, the main focus will be on reporting descriptive statistics. However, to obtain preliminary insights into the intervention effect for future studies, we will perform analyses using an adherence variable and a measurement time point variable while acknowledging the limitations of a single-arm study. To ensure estimation stability and model convergence, the analysis will begin with a simple one: a linear mixed-effects model with random intercepts only, treating measurement time point as a continuous variable. Parameter estimation will be performed using Bayesian statistics. The obtained posterior distribution can serve as the prior distribution for future studies. The posterior probability of the regression coefficient associated with the interaction term of the time and adherence variables being 0 or greater is mainly reported. If no serious estimation problems are found, more complex models, such as ones with random slopes or regarding the time variable as categorical, would also be performed.

### 2.8 Data collection and management

The coordination and data collection and management roles are basically performed by the authors themselves. For MMSE, MoCA, LMs I and II, and verbal fluency tests, we will outsource the work to an external organization and have licensed psychologists conduct the tests. The outsourced psychologists are only informed of participants’ ID, sex, along with years of education (required for scoring the MoCA). Records of other psycho-cognitive measurements and questionnaires, including NCGG-FAT, will be handled by team members registered in the ethics application. Those team members have access to all background information about participants. Those who enter the data obtained may include those who will later be involved in data analysis. Only persons registered in the ethics application documents can access these data.

## 3 Ethical consideration and trial registration

This study was approved by the ethics committee of RIKEN (Ethical approval number: RIKEN-W1-2025-001) and prospectively registered in the University Hospital Medical In-formation Network clinical trial registry (UMIN000057738) prior to the start of recruitment on April 29, 2025 [53]. At the initial registration, only SCO, IRI, ZTPI, and NCGG-FAT were registered as outcomes; however, the latest version, released on August 5, 2025, added other outcomes listed in Table 1. All relevant items from the WHO Trial Registration Data Set are available in the present paper and at [53].

## 4 Discussion

### 4.1 Strength

The practical lessons learned from this long-term exploratory intervention, which relaxes exclusion criteria and potentially includes participants with diverse characteristics such as cognitive decline and limited experience with device use, would provide valuable information to researchers considering implementing conversation-based interventions, digital health approaches, and related studies in community settings. For example, in the previous RCT involving a short-term robot-assisted intervention [26], older adults with a low frequency of digital device use in daily life tended to have fewer utterances. Whether such issues are resolved over the long term is a matter of considerable interest.

Another major feature of this study is the acquisition of a dataset that combines conversational data across various topics with annual cognitive function test scores and questionnaire responses. This dataset can provide valuable information for intervention research. Recently, cognitive interventions that integrate various domains such as diet and exercise, i.e., multidomain intervention, have gained attention for their efficacy [54]. As mentioned in Section 2.6.5, Ref. [17], using conversation data, found that utterance types associated with higher logical memory scores were frequently observed in conversation topics related to outings, and based on this finding, noted the promise of the combination of the Coimagination method and the process of participants walking together through the city taking photos. This can be seen as laying the groundwork for multi-domain interventions that incorporate activities from other domains, such as light physical exercise, into conversation-based interventions. As demonstrated by this example, this dataset is expected to help answer the question of which domains of activities elicit specific conversation patterns that serve as effective cognitive training.

Furthermore, longitudinal observations of outcomes in this study may provide valuable insights into the research fields from which these outcomes originate. For example, there have been studies conducting repeated measurements of ZTPI, but these have involved short-term follow-ups [35], [55], cross-sectional associations with age [56], or have targeted younger populations [35], [55]. In this regard, the observation of ZTPI in older adults over five years in this study would yield insightful information, even though it is conditional on participants being involved in a specific intervention program.

Moreover, Ref. [57] found a nonlinear relationship between SCO and age, particularly the increase in SCO in older age. This result provides an important implication for our study, which indicates that when tracking changes in SCO, we should consider not only the effect of the intervention but also that of age. In contrast to Ref. [57], which conducted between-individual analyses, our study will capture within-individual changes in SCO together with conversation data. These data will contribute to better understanding both intervention and age effects by employing a predictive model that accounts for the time dependence between the adherence variable obtained from conversation data and SCO and incorporates age as an exogenous variable.

### 4.2 Limitations

We have previously conducted RCTs in both Wako [26], [58] and Kishiwada [19], [59]. The recruitments were conducted through the random distribution of 3,000–4,000 letters. Those who have voluntarily participated in our past research in this way are already familiar with our research team and are likely to have a high level of interest in dementia prevention research. In fact, we have sent information about our future activities to those who expressed interest among past participants. Therefore, we cannot rule out the possibility that they may apply to participate in this study again. This means that the proportion of participants who are familiar with our intervention methods is higher than that of ordinary samples, which may lead to an overestimation of feasibility and an earlier appearance of intervention effects than usual. This point must be taken into consideration when interpreting the results. In particular, in Kishiwada, RCTs were conducted from June 2023 to February 2024 and from June 2024 to February 2025 to verify the effectiveness of the Coimagination intervention program (albeit through remote conversations, unlike this study), due to which this impact may be particularly pronounced.

This study plans to measure various outcomes related to the program. Despite this, measurements of confounding factors necessary to properly evaluate the intervention effect on each outcome may be insufficient. Although analytic strategies to mitigate issues arising from this single-arm study have already been described, the issue on confounding factors should be kept in mind. While such a issue may be considered limitations when confined to the exploratory analyses conducted within this protocol, we view them to be a strength of our study, as they enable the efficient conduct of future full-scale studies by selecting important outcomes based on the results of this study and enhancing the measurement of potential confounding factors accordingly.

## Declarations

### Funding

This work is supported by JSPS KAKENHI (Grant numbers: 22H00544 and 25K05587) and JST (Grant number: JPMJPF2101).

## Competing interests

No commercial sponsor is involved. The founders have no role in this study other than providing research funding.

## Availability of data and materials

As this manuscript is a study protocol, no data have been analyzed yet. Upon completion of the study, the datasets are expected to be available upon reasonable request to the authors.

## Authors’ contributions

MOM and TS conceived the study. The first draft was written by TS. KK and MOM developed the intervention system. HS supported the revision of the manuscript. TS, KK, and MOM were involved in participant recruitment activities, including briefing and trial sessions. All authors reviewed and approved the final manuscript. In the conduct of the study, TS and KK will mainly collect data. TS will perform the quantitative analysis. KK will manage the operation and maintenance of the intervention equipment and provide technical support to study participants and the research team during the study. All authors will contribute to the interpretation of the findings. The principal investigator is MOM.

## Acknowledgments

The authors sincerely thank Sachiko Iwata, Kosuke Fukumori, Hiroyuki Taira, Tomoko Suzuki, Naoko Watabe, and the staff members of the Wako and Kishiwada city offices and Nobana Health Promote Co., Ltd. for their invaluable contributions to preparing and conducting this study.

## Notes

### Competing Interest Statement

The authors have declared no competing interest.

### Clinical Trial

UMIN000057738

### Author Declarations

Institutional Review Board of RIKEN gave ethical approval for this work.

